# Prioritizing Cardiovascular Disease-Associated Variants Altering NKX2-5 Binding through an Integrative Computational Approach

**DOI:** 10.1101/2023.09.01.23294951

**Authors:** Edwin G. Peña-Martínez, Diego A. Pomales-Matos, Alejandro Rivera-Madera, Jean L. Messon-Bird, Joshua G. Medina-Feliciano, Leandro Sanabria-Alberto, Adriana C. Barreiro-Rosario, Jessica M. Rodriguez-Rios, José A. Rodríguez-Martínez

## Abstract

Cardiovascular diseases (CVDs) are the leading cause of death worldwide and are heavily influenced by genetic factors. Genome-wide association studies (GWAS) have mapped > 90% of CVD-associated variants within the non-coding genome, which can alter the function of regulatory proteins, like transcription factors (TFs). However, due to the overwhelming number of GWAS single nucleotide polymorphisms (SNPs) (>500,000), prioritizing variants for in vitro analysis remains challenging. In this work, we implemented a computational approach that considers support vector machine (SVM)-based TF binding site classification and cardiac expression quantitative trait loci (eQTL) analysis to identify and prioritize potential CVD-causing SNPs. We identified 1,535 CVD-associated SNPs that occur within human heart footprints/enhancers and 9,309 variants in linkage disequilibrium (LD) with differential gene expression profiles in cardiac tissue. Using hiPSC-CM ChIP-seq data from NKX2-5 and TBX5, two cardiac TFs essential for proper heart development, we trained a large-scale gapped k-mer SVM

(LS-GKM-SVM) predictive model that can identify binding sites altered by CVD-associated SNPs. The computational predictive model was tested by scoring human heart footprints and enhancers in vitro through electrophoretic mobility shift assay (EMSA). Three variants (rs59310144, rs6715570, and rs61872084) were prioritized for in vitro validation based on their eQTL in cardiac tissue and LS-GKM-SVM prediction to alter NKX2-5 DNA binding. All three variants altered NKX2-5 DNA binding. In summary, we present a bioinformatic approach that considers tissue-specific eQTL analysis and SVM-based TF binding site classification to prioritize CVD-associated variants for in vitro experimental analysis.

**Graphical Abstract:** 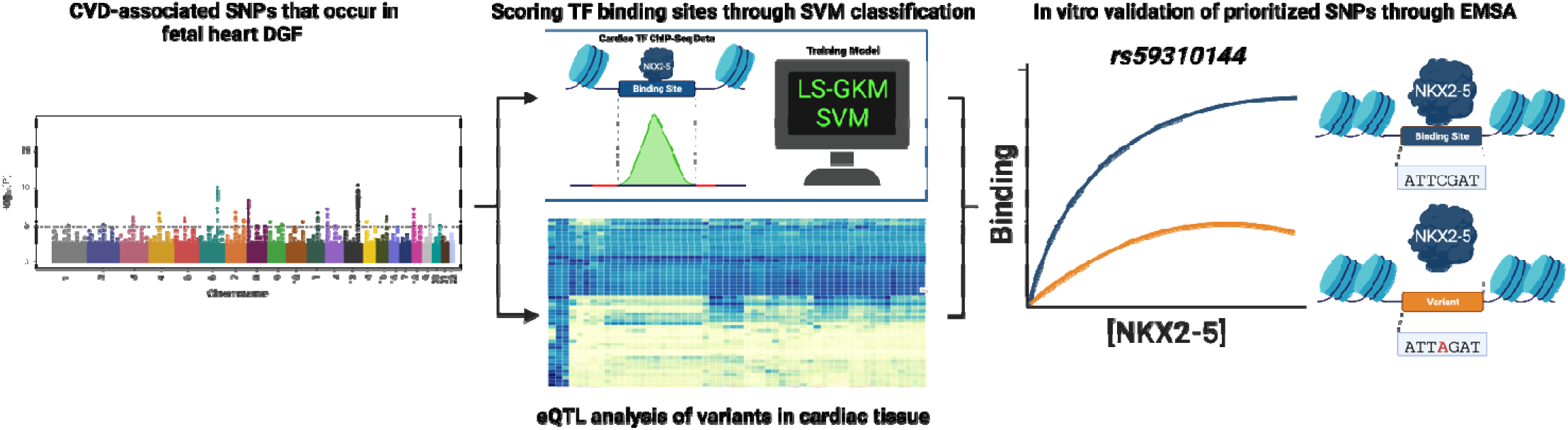

## Introduction

Cardiovascular diseases (CVDs) are the leading cause of death worldwide and encompass multiple disorders (coronary artery disease, congenital heart disease, stroke, etc.), many of which are heritable. ^1–5^ Genome-wide associations studies (GWAS) have mapped over 90% of CVD-associated variants within non-coding regions of the genome (promoters, enhancers, introns, 5⍰/3⍰ UTRs, etc.). ^6,7^ Non-coding single nucleotide polymorphisms (SNPs) can impact phenotype by altering gene regulatory mechanisms, such as transcription factor (TF)-DNA binding and gene expression. ^8–11^ NKX2-5 and TBX5 are cardiac TFs that regulate gene expression in the developing heart. ^12–17^ Previous research has identified CVD-associated SNPs that alter cardiac TF-DNA binding, but further research is required to establish causality. ^18–22^ However, with the overwhelming number of GWAS SNPs (>500,000), prioritizing potential CVD-causing variants for experimental validation remains challenging.

One approach to address this challenge is implementing predictive models to identify variants that create or disrupt TF binding sites (TFBS). ^23–25^ Large-scale gapped k-mer (LS-GKM) support vector machine (SVM) predictive models can be trained to identify TFBS by using in vitro or in vivo DNA-binding data, such as chromatin immunoprecipitation followed by sequencing (ChIP-seq). LS-GKM-SVM models outperform traditional approaches, such as position weight matrix (PWM)-based methods, by considering complex sequence features like dinucleotide interactions, longer/gapped k-mers, and intracellular patterns.^26–29^ LS-GKM-SVM predictive models can be trained with ChIP-seq data from specific cell lines or tissue to integrate relevant epigenomic and regulatory context.^23^

In this work, we present an integrative approach to prioritize functional non-coding variants that can contribute to the biology of CVDs. Using publicly accessible data from the GWAS catalog ^30^, GTEx Portal ^31^, ENCODE ^32^, ChIP-Atlas ^33^, and Remap ^34^, we compiled a list of CVD-associated SNPs linked with a differentially expressed gene in cardiac tissue. We trained a LS-GKM-SVM predictive model with ChIP-seq data from NKX2-5 and TBX5 in human-induced PSC-derived cardiomyocytes (hiPSC-CM). Both models were used to score previously identified heart DNase I hypersensitivity genomic footprints (DGF) ^35^ that colocalize within putative cardiac enhancers ^36^ and tested them through in vitro binding by electrophoretic mobility shift assay (EMSA). Our predictive model was successful at identifying NKX2-5 and TBX5 binding sites and distinguishing between DNA sequences with different binding affinities.

Having validated DGF scored by the predictive model, we scored all CVD-associated SNPs to alter NKX2-5 DNA binding. We chose three variants (rs59310144, rs6715570, and rs61872084) to prioritize for in vitro validation based on their expression quantitative trait loci (eQTL) in cardiac tissue and LS-GKM-SVM prediction to alter NKX2-5 DNA binding. All three variants were validated through EMSA and resulted in changes on NKX2-5 DNA binding. In short, we present a bioinformatic approach that considers tissue-specific eQTL analysis and SVM-based TF binding site classification to prioritize functional CVD-associated SNPs.

## Methods

### Data

ChIP-seq data sets for NKX2-5 and TBX5 from human induced pluripotent stem cell-derived cardiomyocytes (HiPSC-CM) were collected from the ChIP-Atlas ^33^ and Remap ^34^ databases. Dnase I hypersensitivity footprints for fetal heart tissue (left atrium, right ventricle), heart fibroblast, and differentiated cardiomyocytes were obtained from ENCODE (ENCSR764UYH). ^32^Heart enhancers were downloaded from the supplementary files from Dickel et al. ^36^ Disease or trait-associated SNPs were downloaded from the GWAS catalog (gwas_catalog_v1.0-associations_e0_r2022-11-29.tsv).

### Model training

Large-scale gapped k-mer (LS-GKM) was implemented to perform predictions on TF-DNA binding affinity for NKX2-5 and TBX5. ^37,38^ LS-GKM was downloaded through the Comprehensive R Archive Network (CRAN), for Linux, Mac OS, and Windows platforms. For each TF ChIP-seq bed file, peaks were sorted by intensity and the top 1,000 peaks were used as a positive set for training the predictive models. The *genNullSeqs()* function from the gkmSVM package in R was used to generate negative training by selecting unbound sequences of the same length, chromosome, and GC content as the positive training file. The *gkmtrain()* function was used to train the SVM classifiers. The following parameters were used to train the model using a fivefold cross-validation: word length (*l*) = 11 and the number of informative positions (*k*) = 7 (gkmtrain -x 5 -L 11 -k 7 -d 3 -C 1 -t 2 -e 0.005). Model performance was assessed via receiver operator characteristic (ROC) and precision-recall curves (PCR) area under the curve (AUC) using the gkmSVM package in R.

### Sequence Scoring

The models for each TF were used to predict TF-DNA binding through weighted scoring. The gkmpredict() function was used to score 18 bp sequences within 519,540 DGF from cardiac tissue that were found within previously identified human heart enhancers. These sequences were identified by intersecting genomic coordinates of ∼1.6 million DHFs from cardiac tissue with >80,000 putative enhancers active in fetal and adult human hearts that were identified through ChIP-seq. All function parameters were set to their default values and *gkmpredict()* was used to generate an output file listing all sequences and their respective assigned scores by the classifier model for NKX2-5 and TBX5 binding predictions. Positive scores predicted TF-DNA binding, while negative scores predicted no binding activity.

### Motif Extraction from LS-GKM Models

We scored and sorted every possible 11-mers and selected the top 1,000 sequences for the generation of a Position Weight Matrix (PWM) using the Multiple Em for Motif Elicitation (MEME)^39^ web-based tool default parameters to generate a logo.

### Cardiovascular disease-associated risk-variants Identification

Variants from the GWAS catalog were downloaded and filtered to identify CVD or trait-associated SNPs. Variants were filtered from the “DISEASE/TRAIT” column using the following function:

> grepl(‘heart|cardiac|aortic|atrial|ventric|cardio|vascular|artery|coronary|myocardial|valve|cardio|cardium|stroke’, ‘DISEASE/TRAIT’)

CVD SNPs were intersected with human putative enhancers active in the human heart and DGF from the fetal heart. CVD-associated SNPs that occur within human heart enhancers and footprints were expanded to include variants in linkage disequilibrium (LD) using the LDLinkR package. ^40^ CVD-associated SNPs and variants in LD found in cardiac tissue (heart atrial appendage and left ventricle) with differentially expressed genes were identified through the Genome Tissue Expression (GTEx) Portal database.

### NKX2-5 and TBX5 expression and purification

The NKX2-5 homeodomain (HD) gene (Asp16 to Leu96) was cloned in pET-51(+) expression vector containing an N-terminal Strep•Tag II® and a C-terminal 10× His•Tag® through Gibson Cloning and purified through Ni-NTA affinity chromatography, as previously described. ^18^ The human TBX5 gene (Clone ID HsCD00079979, DNASU Plasmid Repository, AZ) was cloned in pEU-E01-GST-TEV-MCS-N1 (Cambridge Isotope Laboratories, Inc. CFS-PEU-V1.0) vectors using Gibson Assembly (New England Biolabs, Inc). Clones were verified by Sanger Sequencing from the University of Wisconsin Biotechnology Center DNA Sequencing Facility. Protein expression was made using the Wheat Germ Cell-Free Protein Expression from the CellFree Sciences Co following the manufacturer’s protocol. Protein expression was confirmed through an SDS-PAGE followed by Western Blot using Anti-GST HRP-conjugated (NB100-63173) antibody (Novus Biological).

### Electrophoretic mobility shift assay

NKX2-5 and TBX5 binding to their respective scored sequences of human heart footprints and enhancers were evaluated using 20 bp sequences that contained an additional 20 bp constant sequence for IR-700 fluorescent marking (IDT). All sequences were ordered in IDT and are available in **Supplementary Table 1**. The IR-700 fluorophore was added to all the sequences through a primer extension reaction and purified using the QIAquick® PCR Purification Kit (Qiagen 28106). Binding reactions were performed in binding buffer (50 mM NaCl, 10 mM Tris-HCl (pH 8.0), and 10% glycerol) and 5 nM fluorescently labeled dsDNA. Five concentration points were employed for purified NKX2-5 HD ranging from 50 nM to 2000 nM. Cell-free TBX5-DNA binding was evaluated using four TBX5 dilutions (1, 1/5, 1/10, and 1/25) of the cell-free extract. Binding reactions were incubated for 30 min at 30°C followed by 30 min at room temperature before loading onto a 6% polyacrylamide gel in 0.5x TBE (89 mM Tris/89 mM boric acid/2 mM EDTA, pH 8.4). The gel was pre-ran at 85 V for 15 min, loaded at 30 V, and resolved at 75 V for 1.5 h at 4°C. Gels were imaged with Azure® Sapphire Bio-molecular Imager with 658 nm excitation and 710 nm emission.

Binding curves were generated by first quantifying the fluorescence signal in each DNA band using ImageJ. Background intensities obtained from blank regions of the gel were subtracted from the band intensities. The fraction of bound DNA was determined using ***Equation 1***. The fraction of bound DNA was plotted versus the TF concentration. Binding curves were obtained by “one-site specific binding” non-linear regression using Prism software.

**Equation 1**. Binding Affinity from the integrated density of bound and unbound bands.

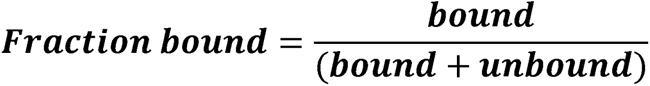

## Results and Discussion

To identify potential CVD-causing SNPs, we downloaded the GWAS catalog and filtered the data to keep cardiovascular disease or trait-associated SNPs (e.g., congenital heart defects, cardiomyocyte differentiation, stroke, arrhythmia, etc.; full list of SNPs in **Supplementary File 1**). We then intersected the CVD-associated SNPs with a catalog of putative fetal and adult heart enhancers and genomic footprints of fetal hearts, resulting in 1,535 genomic variants. The CVD-associated SNP set was expanded to include SNPs in linkage disequilibrium (LD r^2^>0.8) from diverse populations (EUR, AFR, SAS, EAS, and AMR) and resulted in 9,309 unique SNPs occurring in one or more populations. To evaluate the potential of these SNPs to be biologically relevant in cardiovascular biology, we analyzed gene expression patterns in cardiac tissue with the previously identified variants in the GTex portal. We found 636 differentially expressed genes associated with the previously identified SNPs in the heart atrial appendage or left ventricle. The workflow is illustrated in **Figure 1A** and the list of SNPs associated with differentially expressed genes in cardiac tissue is found in **Supplementary File 1**. The distribution of CVD-associated SNPs is not uniform throughout the genome. We identified chromosomes with a higher frequency of CVD-associated SNPs which contain >1,000 variants (chromosomes 1 and 6) and ∼500 (chromosomes 2, 3, 7, 15, 17, and 22), including those in LD (**Figure 1B**). Chromosomes with a high SNP frequency may have variants evenly distributed among them, like chromosomes 1 and 2, while others contain multiple variants in the same (or near) loci, like chromosomes 6, 10, 15, and 22 (**Figure 1C**). This suggests that certain chromosomes, or specific loci, are enriched with CVD-associated SNPs and contribute to the cardiac phenotype. We also analyzed data from the GTEx database to find genes that are differentially expressed in cardiac tissue (heart atrial appendage and left ventricle) containing the identified CVD-associated SNPs or the variants in LD. We identified 25,479 SNP-Gene pairs (636 unique genes) that were significantly differentially expressed in cardiac tissue (**Figure 1D**). Through this approach, we aimed to narrow the extensive list of non-coding variants and identify functional SNPs that contribute to CVD.

**Figure 1:**
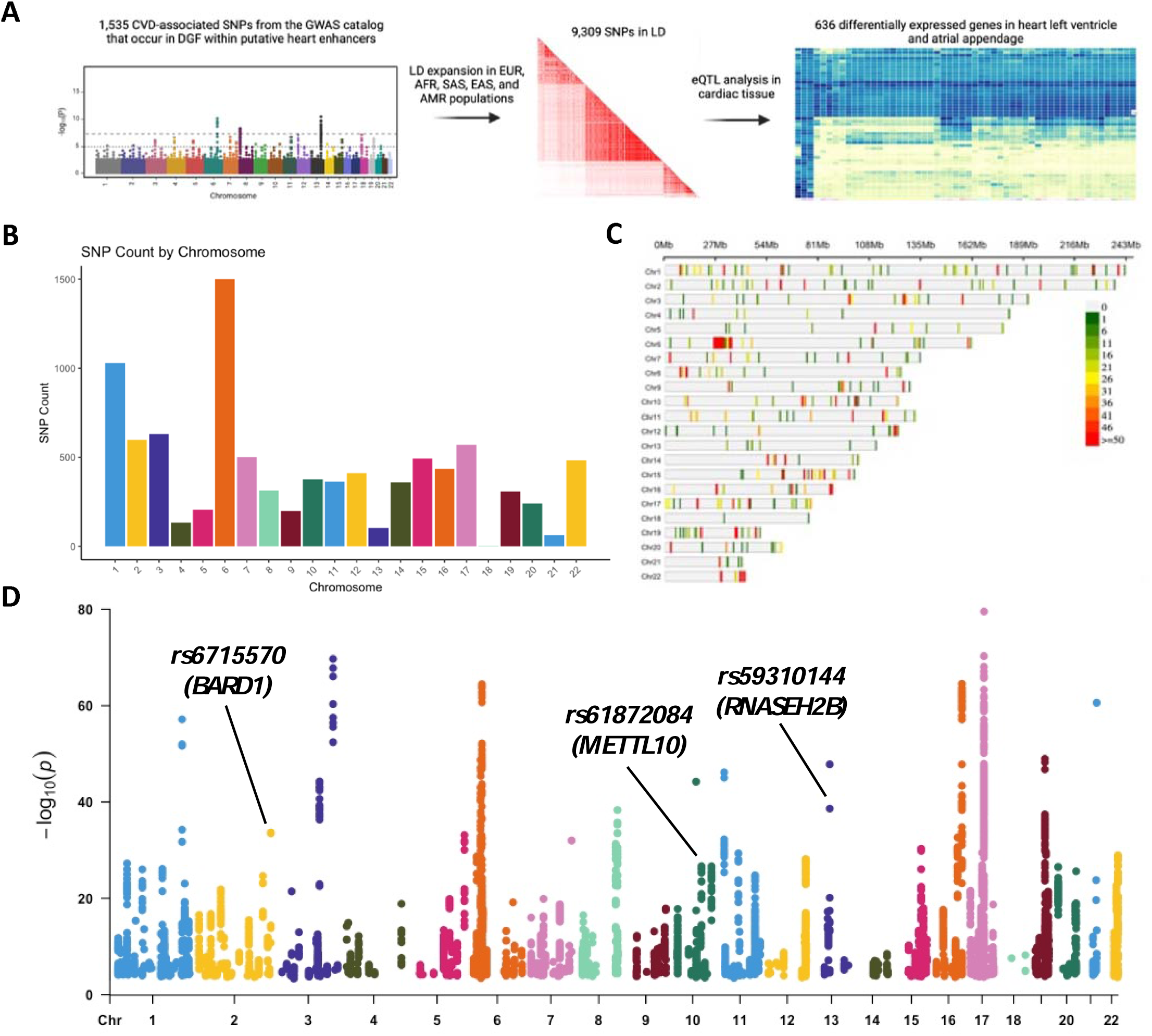
Identification of functional CVD-associated SNPs. **A)** Pipeline to identify potential CVD-causing SNPs. **B)** Number of CVD-associated SNPs per chromosome. **C)** Distribution of SNP frequency within autosomal chromosome, binned by 1Mb windows. **D)** SNP-Gene pairs with differential gene expression in cardiac tissue. Each dot represents a SNP-Gene pair that is differentially expressed in heart atrial appendage or left ventricle in one or more populations. rs6715570-BARD1, rs61872084-METTL10 and rs59310144-RNASEH2B are SNP-Gene pairs that were evaluated in vitro in Figure 3.

We trained a LS-GKM SVM model to prioritize CVD-associated SNPs that alter DNA binding by TFs known to play important roles in heart development and biology. The models were trained using human induced pluripotent stem cell-derived cardiomyocytes (HiPSC-CM) ChIP-seq data for NKX2-5 and TBX5. The 1,000 top-scoring ChIP-seq peaks were used as a positive training set, while unbound sequences of the same length, GC content, and chromosome were used as negative training (**Figure 2A)** The best-performing LS-GKM SVM classifier model trained with NKX2-5 ChIP-seq data (SRX9284027) ^41^ obtained an AUROC value of 0.955 and an AUPRC value of 0.954. The best TBX5 (SRX2023721) ^42^ model obtained an AUROC value of 0.921 and an AUPRC value of 0.912 **(Supplementary Figure 1A-B)**. The models were used to score all possible 2,097,152 non-redundant 11 bp oligomers (11-mers). The 11-mer scores were sorted and the 1,000 top-scoring sequences were used to generate Position Weight Matrix (PWM) using MEME (**Supplementary Figure 1C-D**). The PWMs for both models resulted in DNA binding motifs in agreement with previously described models for NKX2-5 and TBX5. ^43–45^ We proceeded to score ∼520,000 fetal heart DGF that occur heart enhancers to identify genomic loci potentially bound by NKX2-5 or TBX5 (**Figure 2B**). We then chose the DNA sequences with the highest, middle, and lowest scores to test for in vitro binding through EMSA (**Figure 2C, Supplementary Figure 2**). There was agreement between LS-GKM SVM scores and extent of in vitro binding activity for both, NKX2-5 and TBX5. Our results suggest that our LS-GKM SVM model will be able to successfully predict changes in binding affinity between reference and variant DNA sequences that alter cardiac TF-DNA binding.

**Figure 2:**
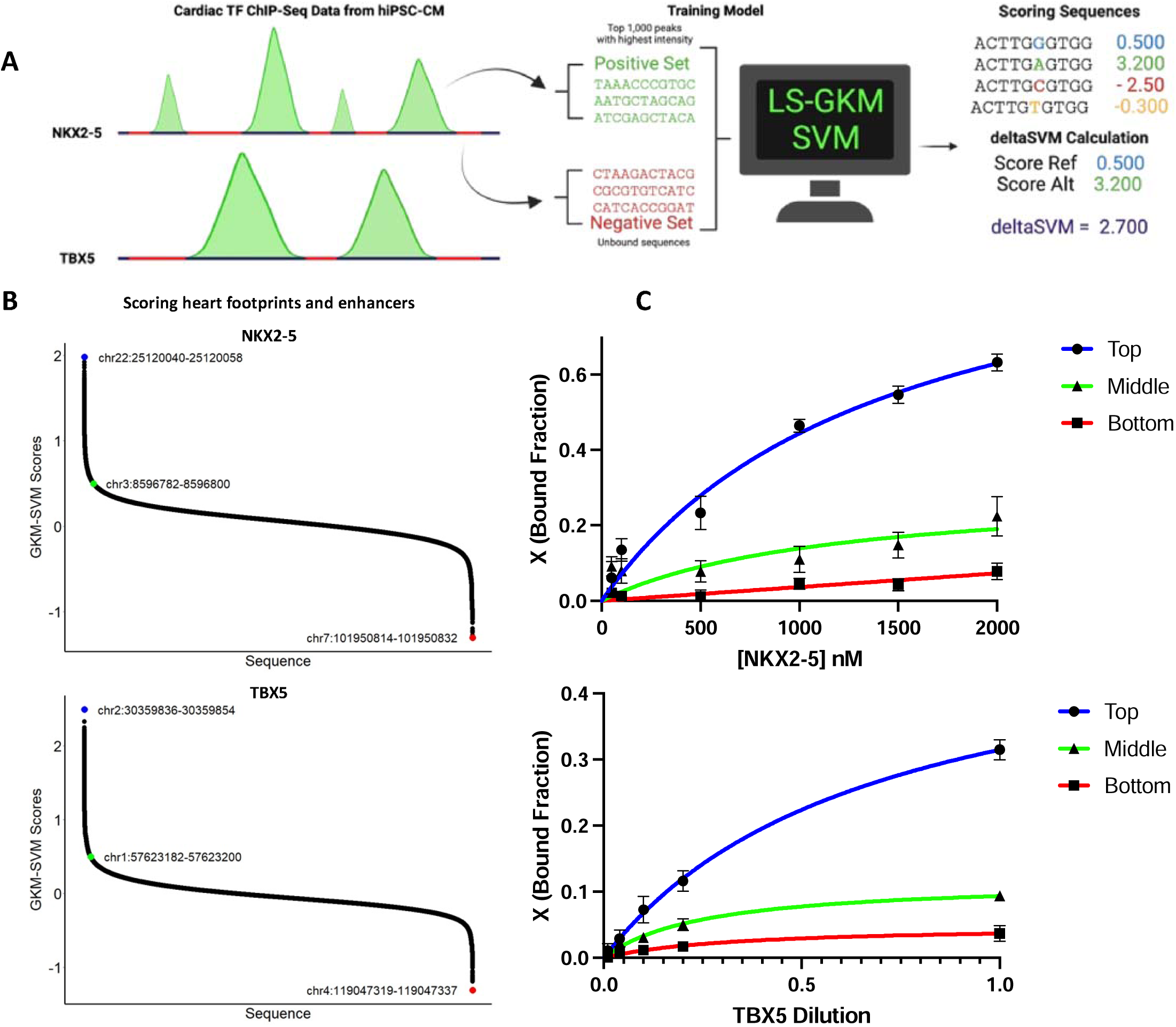
Training and testing of LS-GKM SVM predictive model. **A)** Schematic of model training with NKX2-5 and TBX5 ChIP-seq data from HiPSC-CM. **B)** Scoring of ∼520,000 DGF that occur in heart enhancers with the NKX2-5 (top) and TBX5 (bottom) predictive models. **C)** In vitro testing of predictive model for highest, middle, and lowest scored sequences for NKX2-5 (top) and TBX5 (bottom). For NKX2-5, we tested chr22:25120040-25120058 (circle with blue line), chr3:8596782-8596800 (triangle with green line), and chr7:101950814-101950832 (square with red line). For TBX5, we tested chr2:30359836-30359854 (circle with blue lines), chr1:57623182-57623200 (triangle with green line), and chr4:119047319-119047337 (square with red line).

After successful training and validation of the LS-GKM SVM predictive model, we proceeded to score the 9,309 SNPs to prioritize functional variants. Both reference and alternate allele sequences were scored to predict fold change (deltaSVM score) of TF-DNA binding. We selected three SNPs (rs59310144, rs6715570, and rs61872084) that deltaSVM predicted significant change in NKX2-5 binding and are associated with a differentially expressed gene in cardiac tissue (**Figure 3A, Supplementary Table 2**). When evaluated through EMSA, we observed a differences in NKX2-5 DNA binding between reference and alternate for all three SNPs (**Figure 3B-C, Supplementary Figure 3**). Variants rs59310144 and rs61872084 resulted in a decrease in NKX2-5 DNA binding, while rs6715570 increased binding.

**Figure 3:**
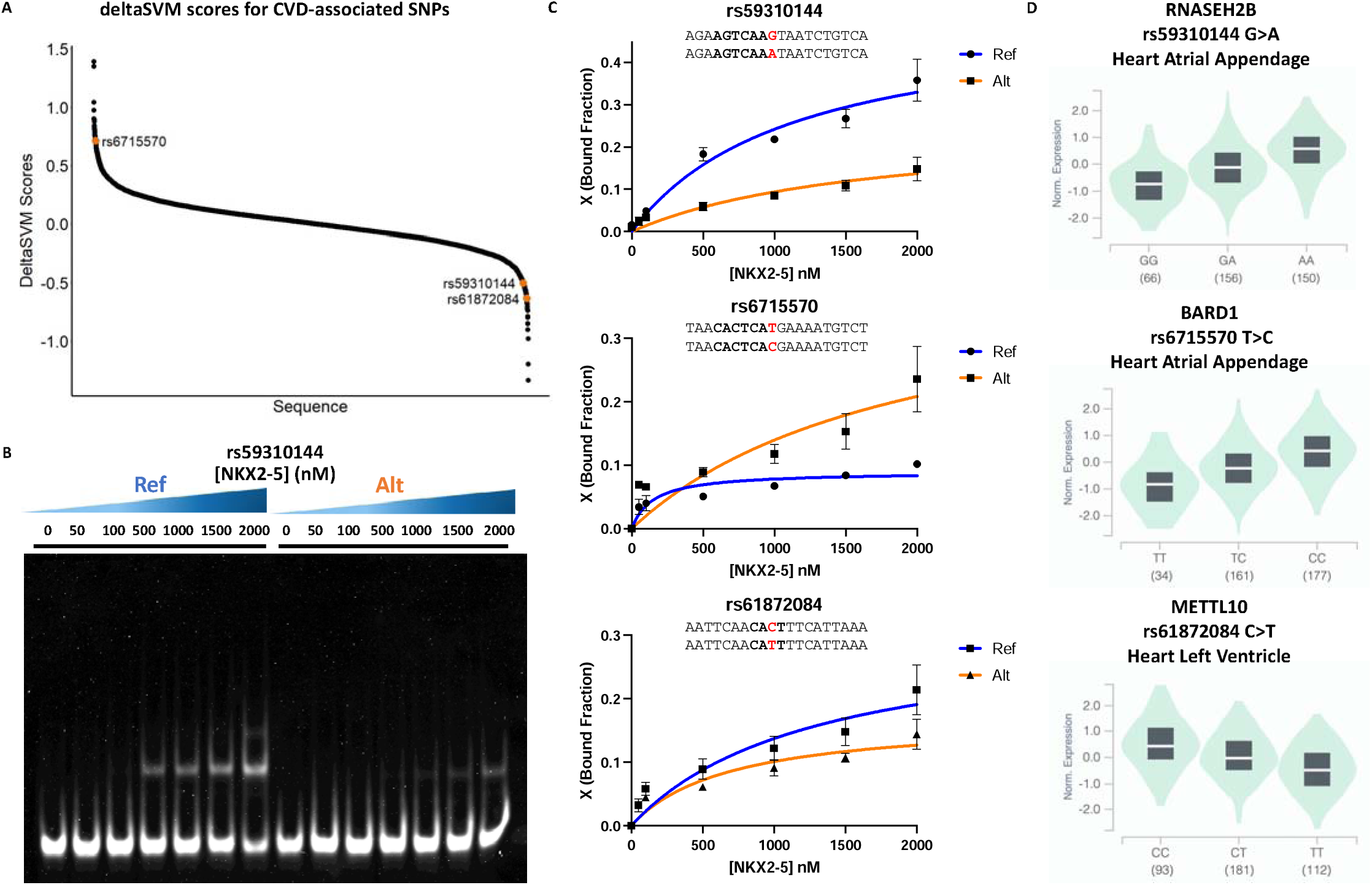
CVD-associated SNPs alter NKX2-5 in vitro binding. **A)** DelstaSVM score distribution of the 9,309 CVD-associated SNPs. **B)** Representative EMSA gel for rs59310144 reference (Ref) and alternate (Alt) alleles. **C)** Binding curves for reference (Ref) and variant (Alt) alleles of rs59310144 (top), rs6715570 (middle), and rs61872084 (bottom). Experiments were performed in triplicates and binding curves show average bound fractio (X) and error bars are standard error. **D)** Cardiac tissue eQTL analysis of *RNASEH2B* (top)*, BARD1* (middle)*, and METTL10* (bottom) expressed in heart atrial appendage or left ventricle when rs59310144, rs6715570, and rs61872084 occur, respectively.

We found that all three SNPs were in eQTLs described in cardiac tissue and identified three genes that are differentially expressed when these variants occur in the heart atrial appendage or left ventricle (**Figure 3D**). RNASEH2B and BARD1 have been previously identified to be differentially expressed in the heart atrial appendage when variants rs59310144 and rs6715570 (respectively) occur. *RNASEH2B*, which has been previously found to be differentially expressed in CVD risk events, is upregulated when the alternate allele of variant rs59310144 is present. ^46^ *BARD1* has also been identified as upregulated when the alternate allele of variant rs6715570 occurs in the heart atrial appendage. Copy number alterations in the *BARD1* locus have been associated with developmental delays, including coarctation of the aorta during early organogenesis and heart development. ^47^ Variant rs61872084 has been identified in the heart’s left ventricle when *METTL10* (Methyltransferase like protein 10) is downregulated when the alternate allele occurs. Accumulation of METTL10 methylated products, such as S-adenosyl-L-methionine, S-adenosyl-L-homocysteine, and homocystein have been correlated with kidney dysfunction and CVD in patients with type 2 diabetes. ^48^ This suggests that NKX2-5 regulation of the *RNASEH2B* (inhibition), *BARD1* (activation), and *METTL10* (activation) genes are possible mechanisms that can be further explored to establish rs59310144, rs6715570, and rs61872084 as causal CVD risk-variants.

## Conclusion

As we continue to research the genetic basis for human disease, the number of identified functional/causal non-coding SNPs continues to grow. Understanding and prioritizing SNPs that contribute to the disease phenotypes is essential. However, we lack a consensus or bioinformatic protocol to prioritize non-coding SNPs that are biologically relevant in the development of human diseases. ^25^ To address this challenge, we applied a GKM-SVM-based model to identify and prioritize potential CVD-causing variants for experimental validation. We leveraged on public data from the GWAS catalog and extracted SNPs that were associated with cardiovascular disease or traits and included variants in LD from multiple populations (EUR, AFR, SAS, EAS, and AMR). These SNPs were analyzed with data from the GTEx database to identify genes that are differentially expressed when these variants are present in cardiac tissue. We tested three SNPs (rs59310144, rs6715570, and rs61872084) associated with a differentially expressed gene (*RNASEH2B*, *BARD1*, and *METTL10* respectively) in cardiac tissue that resulted in changes in NKX2-5 DNA binding activity. Our findings open the possibility that NKX2-5 regulation of *RNASEH2B*, *BARD1*, and *METTL10* is a possible mechanism that can be further researched to determine the causality of CVD-risk variants. Although the etiology of human diseases is complex and multifactorial, this approach can provide crucial information that can be implemented during in vivo experiments or clinical research to address genetic diseases caused by non-coding SNPs. In summary, we believe this bioinformatic approach, which considers tissue-specific eQTL analysis and SVM-based TF binding site classification, is a scalable method that can be applied to multiple types of human diseases.

## Acknowledgments/Funding

This project was supported by NIH-SC1GM127231, NSF [1736026], NSF LSAMP [HRD-2008186], University of Puerto Rico Rio Piedras Institutional Funds (FIPI), Puerto Rico Science, Technology, and Research Trust, and NIH Institutional Development Award (IDeA) INBRE [P20GM103475]. EGPM, DAPM, ACBR, JGMF and JMRR were funded by the NIH RISE Fellowship (5R25GM061151-20). DAPM was funded by NSF [IQ BIOREU 1852259]. EGPM and JMRR were funded by the NSF BioXFEL Fellowship (STC-1231306). ARM and JLMB were funded by NSF HRD-2008186. ARM was funded by NSF REU: PR-CLIMB Program (2050493) and NIH 1T34GM145404-01A1. LSA was funded by NIH ID-GENE Fellowship (1R25HG012702-01). JMRR was funded NSF Graduate Research Fellowship (1744619). Graphical abstract, Figure 1A, and Figure 2A were created in Biorender®.

## Data Availability

All data generated for this study is publicly available at https://github.com/joshuagmedina/cardioDisease_riskVariants (accessed on 29 August 2023).

## Supplementary Material

**Table 1:**
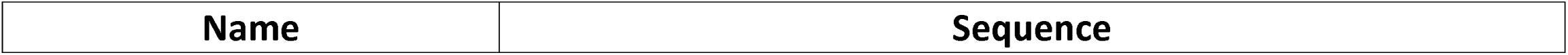
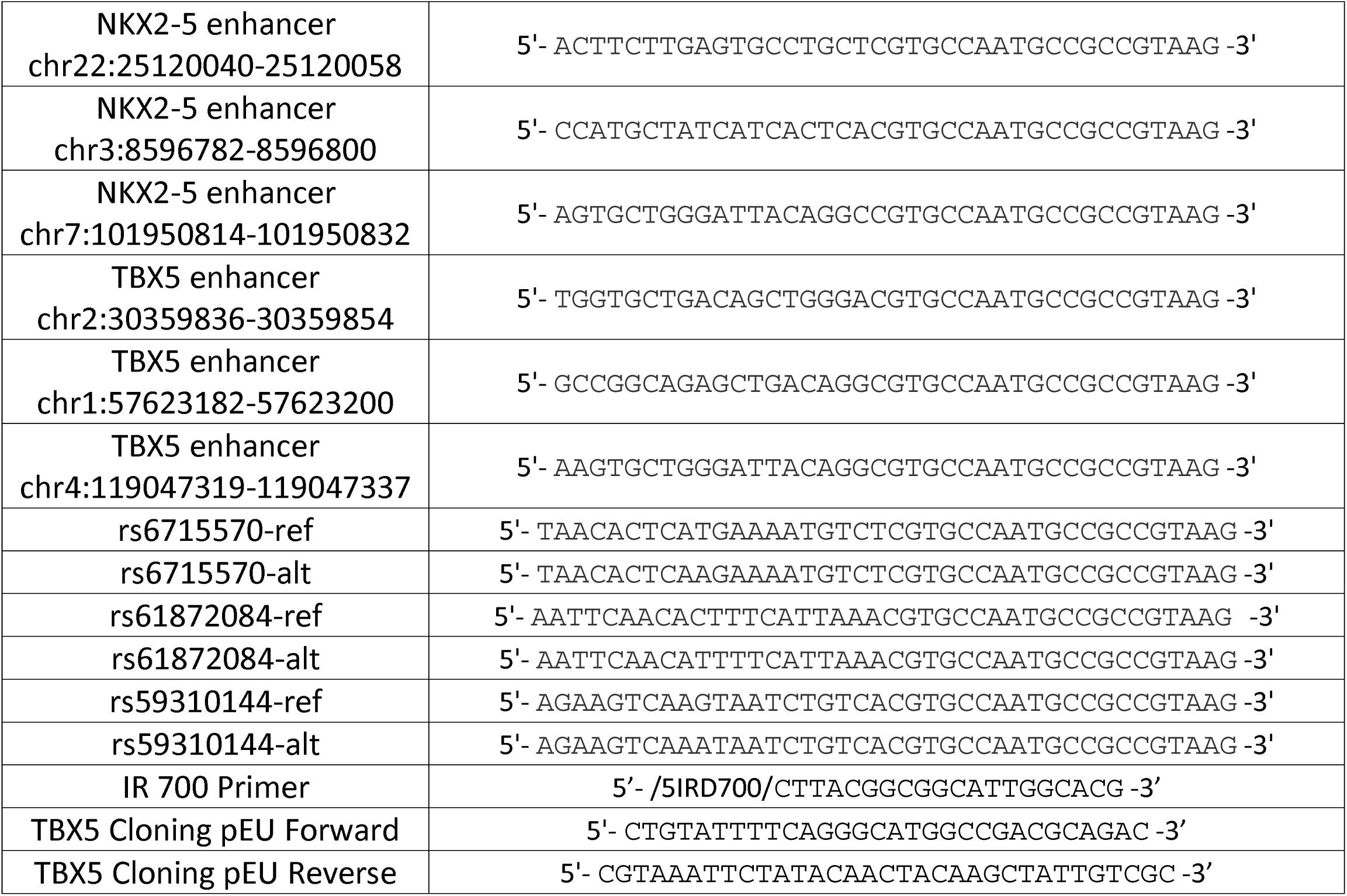
Oligonucleotides used in this work.

**Supplementary Figure 1:**
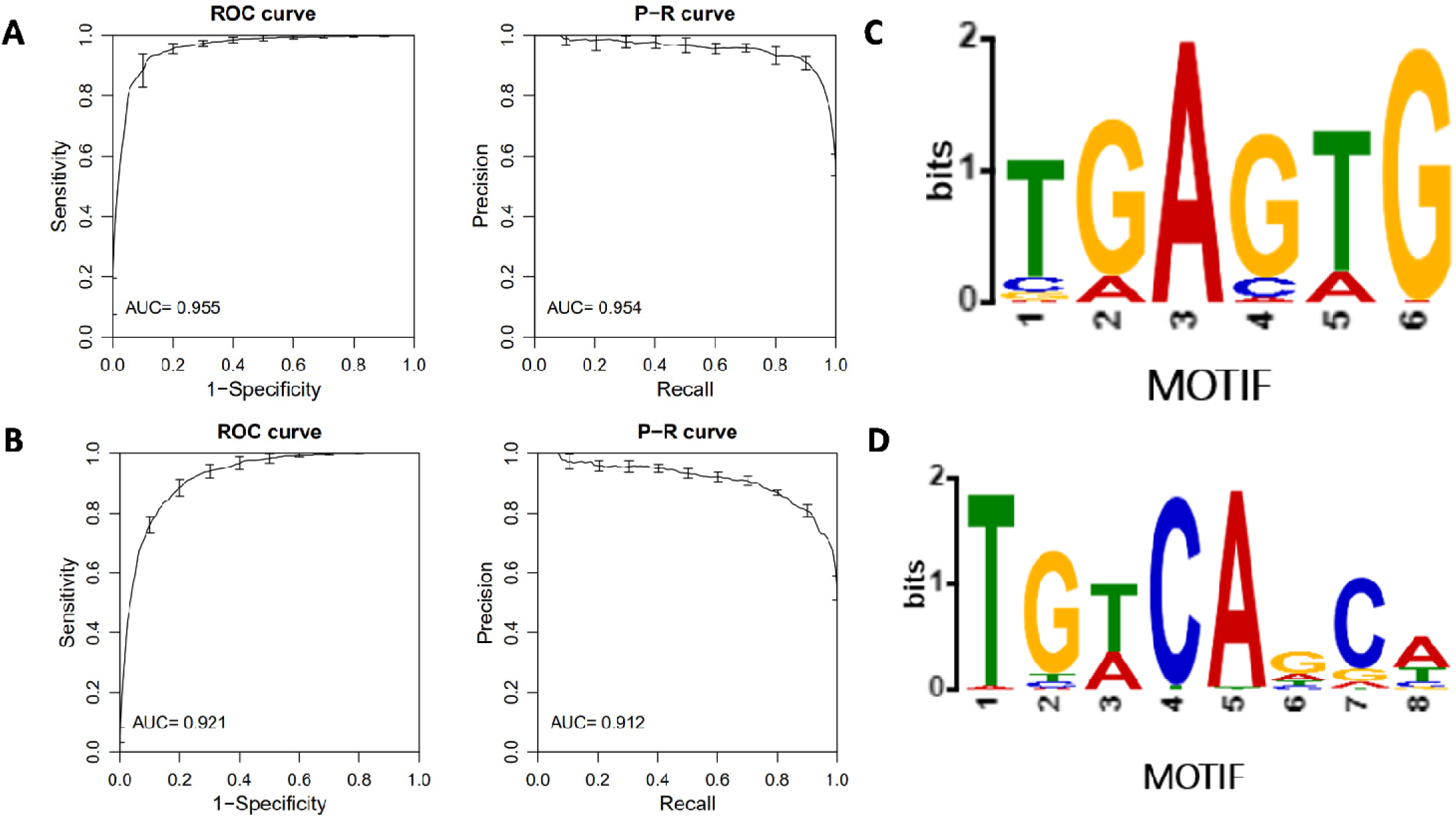
Performance parameters and motif analysis. Performance parameters of **A)** NKX2-5 and **B)** TBX5 as determined by their receiver operating characteristics (ROC) and precision-recall curves. Binding motif for **C)** NKX2-5 and **D)** TBX5 after scoring all possible 11-mers and generating a PWM logo.

**Supplementary Figure 2:**
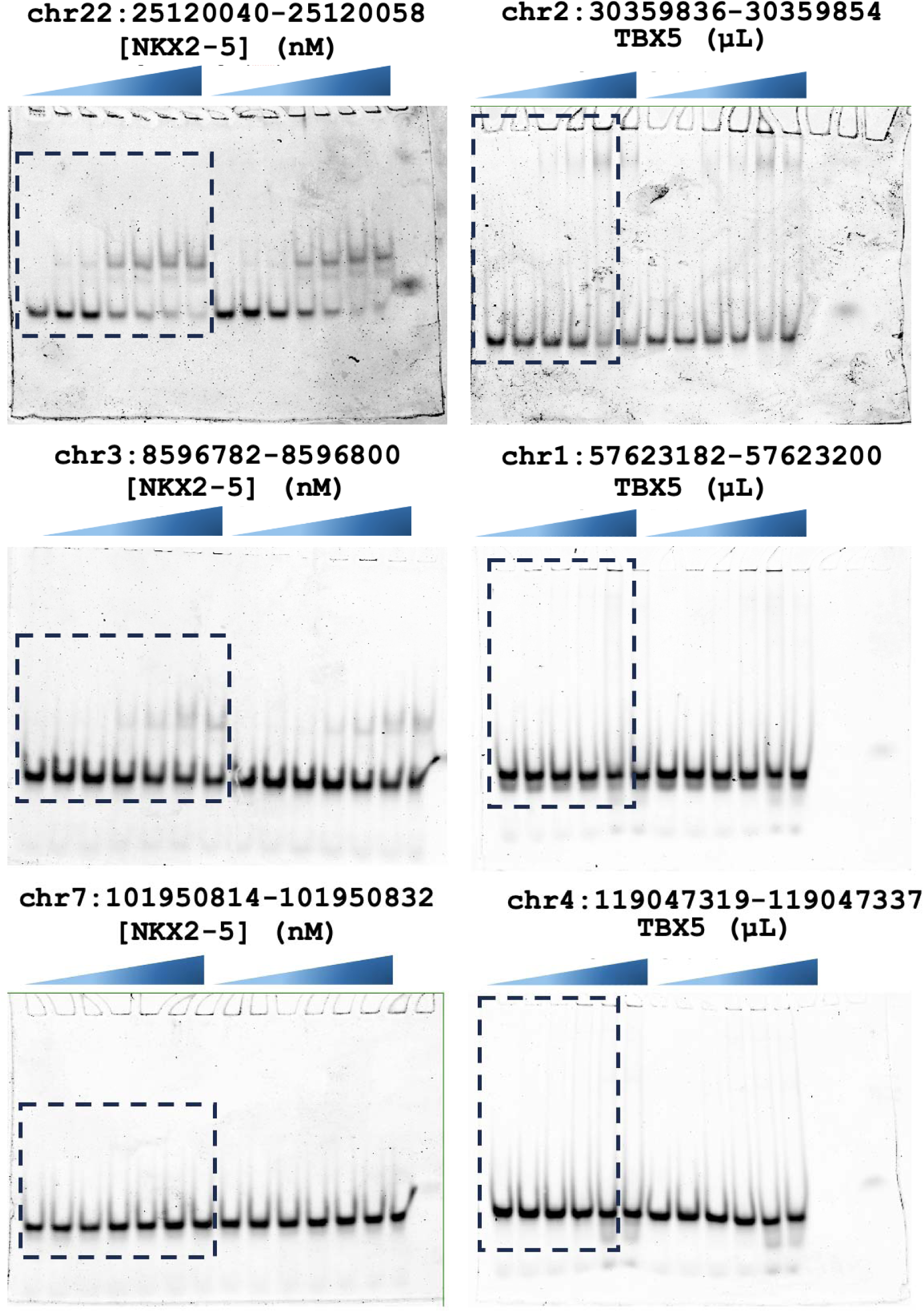
EMSA analysis of heart footprint and enhancers for NKX2-5 (left) and TBX5 (right). All EMSA were performed and triplicates and regions within dashed were used to generate binding curves.

**Supplementary Figure 3:**
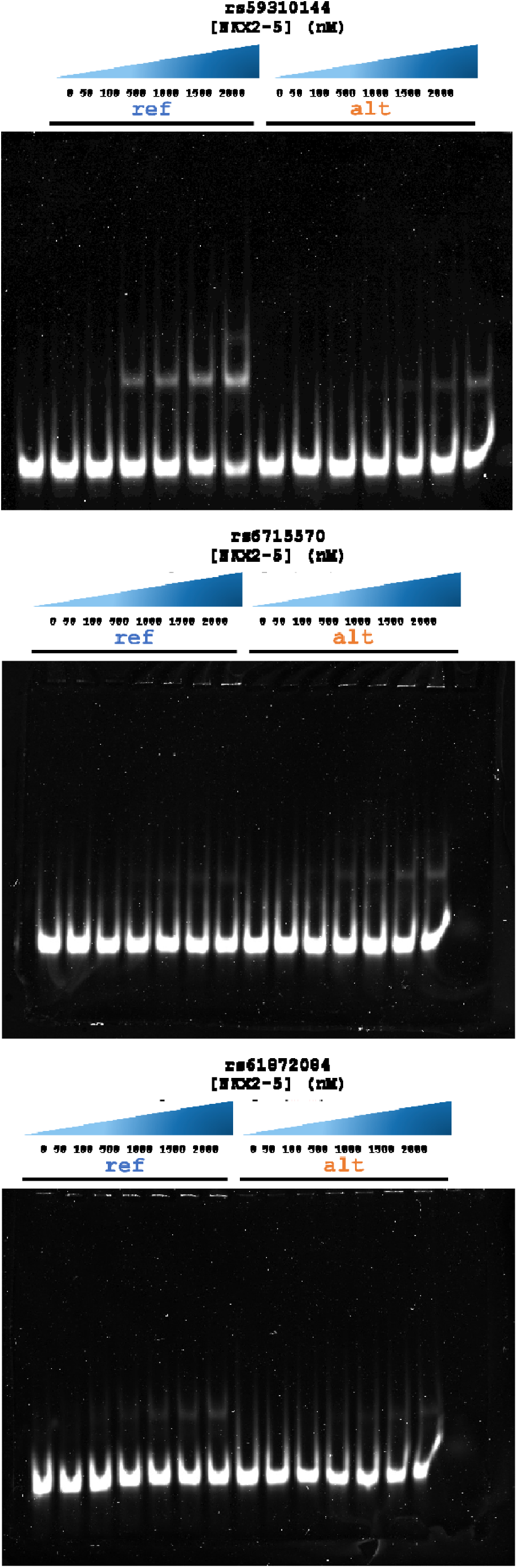
EMSA analysis of three CVD-associated SNPs.

**Supplementary Table 2:** CVD-associated SNPs with differential gene expression in cardiac tissue and predicted impact on NKX2-5 DNA binding.

| Query SNP | LD SNP | Chr | Position | Ref score | Alt score | deltaSVM Score | Gene | Tissue | P value | Association to CVD |
| --- | --- | --- | --- | --- | --- | --- | --- | --- | --- | --- |
| s17074987 | rs59310144 | 13 | 50917644 | G (0.56) | A (0.06) | -0.5 | *RNASEH2B* | Heart - Atrial Appendage | 2.33E-39 | Identified as differentially expressed in CVD risk events |
| rs6435862 | rs6715570 | 2 | 214808716 | T (0.217) | C (0.92) | 0.7 | *BARD1* | Heart - Atrial Appendage | 2.39E-25 | Associated with development delay and coarctation of aorta in early organogenesis and heart development |
| s11245347 | rs61872084 | 10 | 124750493 | C (0.44) | T (-0.2) | -0.63 | *METTL10* | Heart - Left Ventricle | 6.81E-27 | A high concentration of enzymatic product (tHcy) is correlated with kidney dysfunction and CVD |

